# Safety and immunogenicity of SARS-CoV-2 vaccine MVC-COV1901 in adolescents in Taiwan: A double-blind, randomized, placebo-controlled phase 2 trial

**DOI:** 10.1101/2022.03.14.22272325

**Authors:** Luke Tzu-Chi Liu, Cheng-Hsun Chiu, Nan-Chang Chiu, Boon-Fatt Tan, Chien-Yu Lin, Hao-Yuan Cheng, Meei-Yun Lin, Chia-En Lien, Charles Chen, Li-Min Huang

**Author notes:** Co-corresponding authors: Li-Min Huang, Charles Chen.

## Abstract

**Background:** MVC-COV1901 is a subunit SARS-CoV-2 vaccine based on the prefusion spike protein S-2P and adjuvanted with CpG 1018 and aluminum hydroxide. Although MVC-COV1901 has been licensed for emergency use for adults in Taiwan, the safety and immunogenicity of MVC-COV1901 in adolescents remained unknown. As young people play an important role in SARS-CoV-2 transmission and epidemiology, a vaccine approved for adolescents and eventually, children, will be important in mitigating the COVID-19 pandemic.

**Methods:** This study is a prospective, double-blind, multi-center phase 2 trial evaluating the safety, tolerability and immunogenicity of two doses of the SARS-CoV-2 vaccine MVC-COV1901 in adolescents. Healthy adolescents from age of 12 to 17 years were recruited and randomly assigned (6:1) to receive two intramuscular doses of either MVC-COV1901 or placebo at 28 days apart. The primary outcomes were safety and immunogenicity from the day of first vaccination (Day 1) to 28 days after the second vaccination (Day 57), and immunogenicity of MVC COV1901 in adolescents as compared to young adult vaccinees in terms of neutralizing antibody titers and seroconversion rate. The secondary outcomes were safety and immunogenicity of MVC-COV1901 as compared to placebo in adolescents in terms of immunoglobulin titers and neutralizing antibody titers over the study period.

**Results:** Between July 21, 2021 and December 22, 2021, a total of 399 adolescent participants were included for safety evaluation after enrollment to receive at least one dose of either MVC-COV1901 (N=341) or placebo (N=58). Of these, 334 and 46 participants went on to receive two doses of either MVC-COV1901 or placebo, respectively, and were included in the per protocol set (PPS) for immunogenicity analysis. Adverse events were mostly mild and were similar in MVC-COV1901 and placebo groups. The most commonly reported adverse events were pain/tenderness and malaise/fatigue. All immunogenicity endpoints in the adolescent group were non-inferior to the endpoints seen in the young adult and placebo groups.

**Conclusions:** The safety and immunogenicity data presented here showed that MVC-COV1901 has similar safety profile and non-inferior immunogenicity in adolescents compared to young adults.

**ClinicalTrials.gov registration:** NCT04951388.

## Introduction

SARS-CoV-2 is the causative viral agent of COVID-19, an ongoing worldwide outbreak of pneumonia-like respiratory disease [1]. As of early February 2022, close to 400 million confirmed cases and almost 6 million deaths due to COVID-19 have been reported globally [2]. It is now clear that COVID-19 may become an endemic disease [3]. One key reason is the rate of natural viral evolution that is probably too high to permit the extinction of existing viral variants by antiviral treatments before new variants arise [4]. Consequently, existing treatments may have to be regularly redesigned to ensure their ability to optimally interact with and suppress new variants with mutations that reduce efficacy. This issue has recently been highlighted by the Omicron variant which is more infectious than the prior Delta variant and therefore more difficult to eradicate [5]. In addition, this variant contains a new constellation of mutations that reduces efficacy of at least some current treatments [6, 7].

There are also non-scientific reasons that complicate the eradication of COVID-19 and they chiefly include vaccine hesitancy/resistance and the fact that approvals of vaccines for adolescents have lagged approvals for adults because of the increased safety requirements for adolescents that lengthen clinical trial timelines [8]. This issue is now of more importance than ever because the omicron variant appears to be particularly infectious in younger populations [9]. While adolescents generally experience milder symptoms, the lingering sequelae of long COVID can be devastating to their health and development [10]. To this date, the Pfizer-BioNTech and Moderna vaccines are the only widely used vaccines for adolescents and children and these approvals have come only recently [11]. As a result, there remains a lag between vaccination rates in younger people versus adults and this can be a critical gap in COVID-19 transmission control. For example, as of February 2022 in the US, only 56.9% of adolescents aged 12-17 are fully vaccinated compared to 74.6% of adults [12]. Similarly in Europe, 80% of adults are fully vaccinated compared to 70.9% of adolescents aged 15-17 and 35.5% of 10-14 years olds [13]. Therefore, the need to address adolescent vaccination is especially crucial in developing and less-developed countries to address vaccine inequality and disrupt virus transmission.

MVC-COV1901 is a protein-based subunit vaccine comprising S-2P protein, a prefusion stable form of the spike protein of SARS-CoV-2 and toll-like receptor 9 agonist CpG 1018 and aluminum hydroxide as adjuvants [14]. MVC-COV1901 has been approved for emergency use authorization (EUA) in Taiwan for the prevention of COVID-19 in adults above age of 18 [11, 15]. However, safety and immunogenicity of MVC-COV1901 in population under the age of 18 has yet to be explored. To address this unmet need, the phase 2 trial described here explored the safety profile of two doses of MVC-COV1901 in adolescents 12 to 17 years of age and immunogenicity of MVC-COV1901 in adolescents compared to young adults 20-30 years old.

## Methods

### Study design and participants

This is an interim analysis of an ongoing phase 2, prospective, double-blind (investigator/site staff and participants), and multi-center study to assess the safety and immunogenicity of MVC-COV1901 in adolescents aged 12 to 17 years. The study sites included five hospitals/medical centers in Taiwan. Eligible participants were those aged 12 (inclusive) to 17 (not legible if the participant turned 18 years old) at the time of randomization, with body mass index (BMI) at or above the third percentile according to World Health Organization (WHO) at the screening visit, and lack of travel within 14 days of screening and lack of any oversea travelling throughout the study period. Finally, participant and/or the participant’s legal representative must have provided written informed consent. Additional criteria applied to females only: negative pregnancy test, non-childbearing potential or, if with childbearing potential, abstinence or agreement to use medically effective contraception from 14 days before screening to 30 days following the last injection of study intervention. A full list of inclusion and exclusion criteria can be found in the study protocol in the Appendix.

The trial protocol and informed consent form were approved by Taiwan Food and Drug Administration (TFDA) and the ethics committees at the conducting sites: Mackay Memorial Hospital Hsinchu (Hsinchu City), Chang-Gung Memorial Hospital Linkou (New Taipei City), Mackay Memorial Hospital (Taipei City), National Taiwan University Hospital Hsinchu (Hsinchu City), and National Taiwan University Hospital (Taipei City). An Independent Data Monitoring Committee (IDMC) was established to monitor data safety and trial conduct. The study complies with the protocol and statistical analysis plan. This trial was conducted in accordance with the principles of the Declaration of Helsinki and Good Clinical Practice (GCP) guidelines.

### Randomization and blinding

All eligible participants were randomized to receive either MVC-COV1901 or placebo in a 6:1 ratio. All participants were centrally assigned to randomized study intervention using an interactive web response system (IWRS). Before the study was initiated, the log-in information and directions for the IWRS were provided to each site.

This was a double-blind study in which participants and investigators were blinded to study intervention. The IWRS was programmed with blind-breaking instructions and in case of an emergency, the investigator had the sole responsibility for determining if unblinding of participants’ intervention assignment was warranted. For the interim analysis, an independent, unblinded team of the CRO consisting of personnel representing relevant functions, including statistics, programming, and report writing, was involved in the activity of the interim analysis. All activities of the unblinded team were separated from the main blinded team of the study. The Sponsor was blinded until the time of the interim analysis.

As MVC-COV1901 and placebo were visually distinct in their color of appearance, the investigator had assigned unblinded qualified personnel who were not involved in any other aspect of the study to handle the preparation, dispensing, administration, and accountability of the study intervention. Study-specific training was provided to the study site to ensure treatment blinding of all other study staff and the participants.

### Procedures

The study schedule is outlined as in Figure S1. The participants were intramuscularly administered in the deltoid region with two doses of either MVC-COV1901 or placebo (saline) on Day 1 (Visit 2) and Day 29 (Visit 4). The vaccine MVC-COV1901 and placebo were produced at Medigen Vaccine Biologics Zhubei facility (Hsinchu County, Taiwan) in compliance with current good manufacturing practices. MVC-COV1901 contained 15 μg S-2P, 750 μg CpG 1018, and 375 μg aluminum hydroxide at a final volume of 0.5 mL per dose. The placebo contained 0.5 mL of saline solution per dose. Participant samples were collected during six on-site visits: on Days – 28 to 0 (screening Visit), 1 (first vaccination), 29 (second vaccination), 57, 119, and 209. Safety data was monitored by three telephone calls on Days 8, 36, and 85.

For safety analysis, vital signs were assessed before and after each injection. Participants were observed for at least 30 minutes after each injection to identify any immediate adverse events. All participants who received at least one of the two doses of MVC-COV19 scheduled for Day 1 and Day 29 were evaluated for safety up to Day 119 (Visit 8) by assessing the following endpoints: solicited local adverse events (AEs) (up to 7 days after each dose of study intervention), solicited systemic AEs (up to 7 days after each dose of study intervention), unsolicited AEs (up to 28 days after each dose of study intervention), AEs of special interest (AESI), vaccine-associated enhanced disease (VAED) and serious adverse events (SAEs). Solicited AEs were defined as AEs which occurred within 7 days after each dose of study intervention, including local events: pain/tenderness, erythema/redness, and induration/swelling; systemic events: fever, malaise/fatigue, myalgia, headache, nausea/vomiting, and diarrhea. Unsolicited AEs are defined as any untoward medical events other than solicited AEs which occurred within 28 days after each dose of study intervention. The intensities of solicited and unsolicited AEs were graded using grading scales modified from the US FDA Guidance for Industry [16].

To evaluate immunogenicity, live SARS-CoV-2 neutralization assay and anti-SARS-CoV-2 spike immunoglobulin (IgG) ELISA were used as previously reported [15, 17]. Briefly, serially two-fold diluted sera were mixed with an equal volume of SARS-CoV-2 virus (hCoV-19/Taiwan/4/2020, GISAID accession ID: EPI_ISL_411927). The serum-virus mixture was incubated and then added to the plates containing Vero E6 cells, followed by further incubation. The neutralizing antibody titer was defined as the reciprocal of the highest dilution capable of inhibiting 50% of the cytopathic effect (50% inhibiting dilution, ID_50_), which was calculated using the Reed-Muench method. For anti-SARS-CoV-2 spike IgG ELISA, antigen-specific immunoglobulin titers to S-2P protein were evaluated in serum samples collected from participants. The detection and characterization of antigen-specific immunoglobulin were performed by a central laboratory using a validated enzyme-linked immunosorbent assay (ELISA) method with plates coated with S-2P proteins [17].

For standardization of results, the geometric mean titers (GMTs) of ID_50_ from the neutralization assay were converted to International Units (IUs/mL) in the following transformations defined experimentally: y = 1.5001 · x^0.8745^ for the adolescent data and y = 0.4964 · x^1.0334^ for the young adult data, where y is the value of IU/mL and x is the value of the GMT. Similarly, the GMTs from the ELISA assay were converted to Binding Antibody Unit (BAUs/mL) as previously established by multiplying by a factor of 0.0912 [15].

### Outcomes

The primary safety outcome included the occurrence rate of solicited (local and systemic) AEs, unsolicited AEs, AESI, VAED, and SAE from Visit 2 (Day 1) to Visit 6 (28 days after second dose of study intervention). The primary immunogenicity outcomes were neutralizing antibody titers and seroconversion rate (SCR) against live SARS-CoV-2 virus of MVC-COV1901 in adolescents as compared to young adult vaccinees at Visit 6 (Day 57).

The secondary safety outcome included the occurrence rate of ≥ Grade 3 AE, AESI, VAED, and SAE over the whole study period i.e. from Day 1 to 180 days after the second vaccination (Day 209). For secondary immunogenicity outcomes, comparisons of MVC-COV1901 against placebo were performed by measuring and expressing antigen-specific immunoglobulin titers and neutralizing antibody titers in samples taken at Visit 4 (28 days after the first dose of study intervention), Visit 6 (28 days after the second dose of study intervention), Visit 8 (90 days after the second dose of study intervention) and Visit 9 (209 days after the second dose of study intervention).

### Statistical analysis

Three hundred and ninety-nine participants were randomly assigned to study intervention. The sample size was powered to demonstrate that the immunogenicity of MVC-COV1901 in adolescents is non-inferior to that in young adults assessed by neutralizing antibody GMT and seroconversion rate (SCR) 28 days after second dose of study intervention. Demographical and immunogenicity data of young adults (20-30 years old, who were seronegative for SARS-CoV-2 neutralizing antibody at baseline and received MVC-COV1901), were selected from a previous phase 2 trial for comparison [15].

Safety was analyzed with the safety set population, which consisted of all randomized participants who received at least one dose of study intervention. Immunogenicity was analyzed with the per protocol set (PPS) population, which consisted of the individuals who received the planned doses of randomized study intervention per schedule, were seronegative at baseline (neutralizing antibody titer < lower limit of detection at Visit 2), anti-N antibodies negative at Visit 2 and Visit 6, and did not have a major protocol deviations that were judged to impact the critical or key study data.

Seroconversion was defined as at least 4-fold increase of post-study intervention antibody titers from the baseline titer or from half of the lower limit of detection (LoD) if undetectable at baseline. Immunogenicity results were presented as GMTs, GMT ratios, and SCRs with two-sided 95% CIs.

The GMT ratio and two-sided 95% CIs were calculated by exponentiating the mean difference of the logarithms of the titers (the adolescent group cohort minus the young adult group) and the corresponding confidence intervals (based on the two samples t-test). For SCR, chi-square test was used to compare between the two treatment arms.

Immunogenicity primary endpoints were the non-inferiority of neutralizing antibody GMT ratio and SCR at 28 days after second dose of MVC-COV1901 in adolescents compared to young adults. The adolescent group was claimed as non-inferior to young adult group in GMT treatment ratio when the lower bound of the two-sided 95% CI of GMT treatment ratio is greater or equal to 0.67. The adolescent group was deemed as non-inferior to young adults group in SCR treatment difference when the lower bound of the two-sided 95% CI of SCR treatment difference is greater or equal to -10%

The secondary immunogenicity endpoints of study intervention included the GMT, SCR, and GMT ratio of antigen-specific immunoglobulin titers and neutralizing antibody titers at Visit 4 (28 days after the first dose of study intervention), Visit 6 (28 days after the second dose of study intervention), Visit 8 (90 days after the second dose of study intervention) and Visit 9 (180 days after the second dose of study intervention). Only the results at Visit 4 and Visit 6 were available for the interim results. GMT ratio in secondary immunogenicity endpoint was defined as geometric mean of fold increase of post-study intervention titers over the baseline titers.

### Funding and role of funding source

This study was funded by Medigen Vaccine Biologics Corporation (MVC, the study sponsor) and Taiwan Centers for Disease Control (CDC), Ministry of Health and Welfare. MVC had a role in study design, data analysis, and data interpretation, but had no role in data collection, or writing of the clinical report. Taiwan CDC of the Ministry of Health and Welfare had no role in study design, data collection, data analysis, data interpretation, or writing of the report.

## Results

Between July 21, 2021 and December 22, 2021, a total of 405 adolescents were screened and 399 of which were deemed eligible and randomized at 6:1 ratio to receive either MVC-COV1901 (N=341) or placebo (N=58) (Figure 1). To evaluate immunogenicity of vaccination with MVC-COV1901 in adolescents of age 12 to 17 years, immunogenicity parameters were compared between this age group and young adults of age 20-30 years. The demographics of the two age groups are summarized in Tables 1 and S1 and illustrate the absence of any relevant bias between both age groups and between MVC-COV1901 and placebo groups.

**Figure 1.**
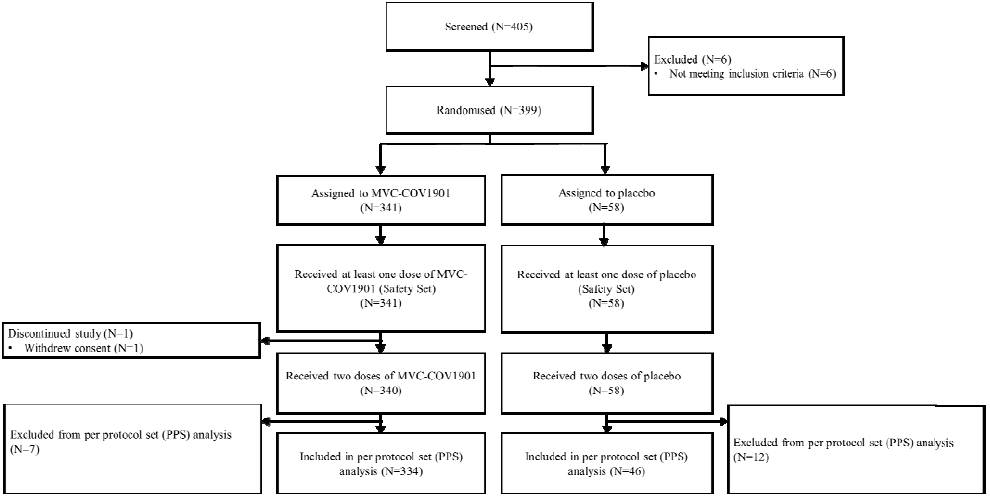
CONSORT Flow diagram of the study.

**Table 1.**
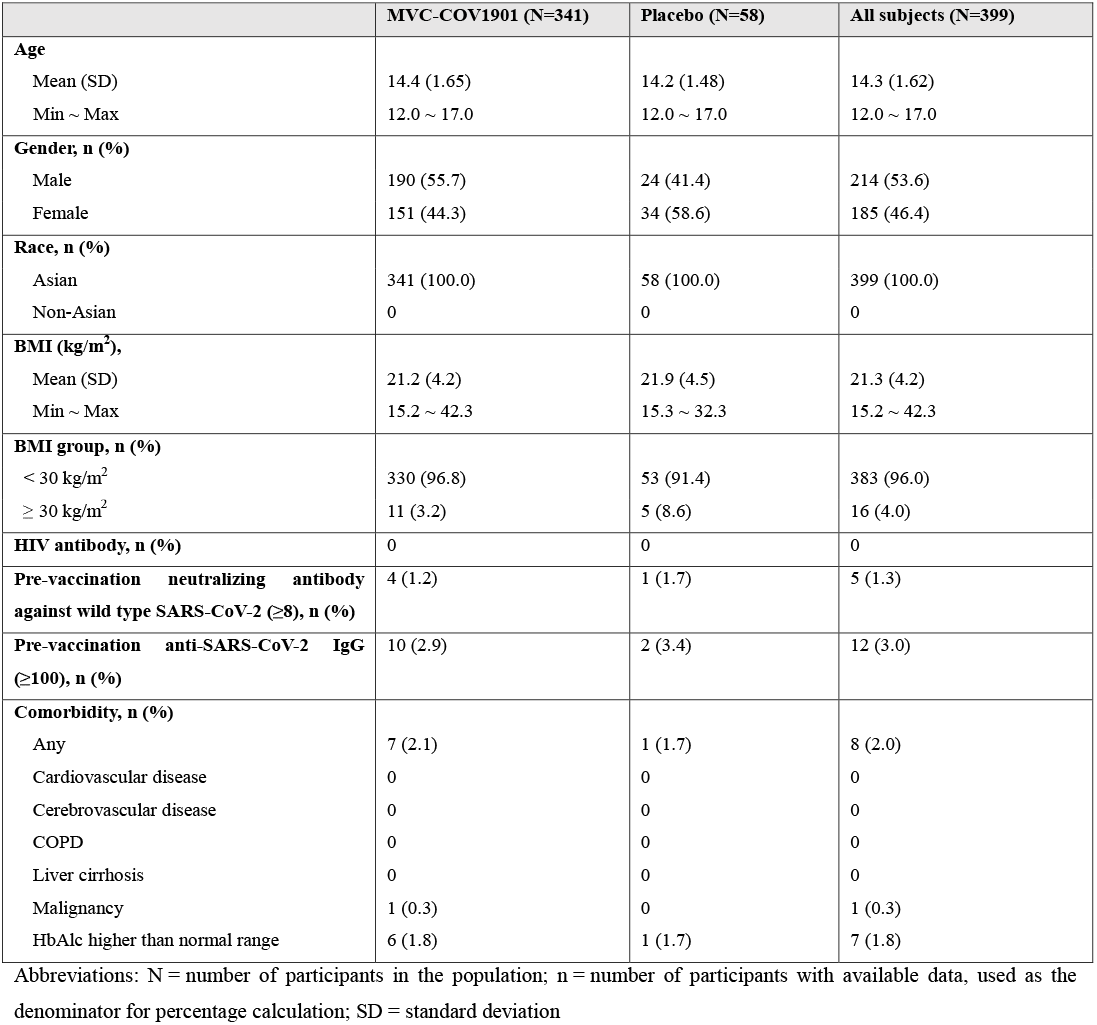
Summary of Demographics of the Participants who Received Study Intervention (Safety Set)

|  | MVC-COV1901 (N=341) | Placebo (N=58) | All subjects (N=399) |
| --- | --- | --- | --- |
| **Age** |  |  |  |
| Mean (SD) | 14.4 (1.65) | 14.2 (1.48) | 14.3 (1.62) |
| Min ~ Max | 12.0 ~ 17.0 | 12.0 ~ 17.0 | 12.0 ~ 17.0 |
| **Gender, n (%)** |  |  |  |
| Male | 190 (55.7) | 24 (41.4) | 214 (53.6) |
| Female | 151 (44.3) | 34 (58.6) | 185 (46.4) |
| **Race, n (%)** |  |  |  |
| Asian | 341 (100.0) | 58 (100.0) | 399 (100.0) |
| Non-Asian | 0 | 0 | 0 |
| **BMI (kg/m<sup>2</sup>),** |  |  |  |
| Mean (SD) | 21.2 (4.2) | 21.9 (4.5) | 21.3 (4.2) |
| Min ~ Max | 15.2 ~ 42.3 | 15.3 ~ 32.3 | 15.2 ~ 42.3 |
| **BMI group, n (%)** |  |  |  |
| < 30 kg/m<sup>2</sup> | 330 (96.8) | 53 (91.4) | 383 (96.0) |
| ≥ 30 kg/m<sup>2</sup> | 11 (3.2) | 5 (8.6) | 16 (4.0) |
| **HIV antibody, n (%)** | 0 | 0 | 0 |
| **Pre-vaccination neutralizing antibody against wild type SARS-CoV-2 (≥8), n (%)** | 4 (1.2) | 1 (1.7) | 5 (1.3) |
| **Pre-vaccination anti-SARS-CoV-2 IgG (≥100), n (%)** | 10 (2.9) | 2 (3.4) | 12 (3.0) |
| **Comorbidity, n (%)** |  |  |  |
| Any | 7 (2.1) | 1 (1.7) | 8 (2.0) |
| Cardiovascular disease | 0 | 0 | 0 |
| Cerebrovascular disease | 0 | 0 | 0 |
| COPD | 0 | 0 | 0 |
| Liver cirrhosis | 0 | 0 | 0 |
| Malignancy | 1 (0.3) | 0 | 1 (0.3) |
| HbA1c higher than normal range | 6 (1.8) | 1 (1.7) | 7 (1.8) |

Abbreviations: N = number of participants in the population; n = number of participants with available data, used as the denominator for percentage calculation; SD = standard deviation

All individuals who received at least one of the two vaccinations were evaluated for safety analysis. The incidences of solicited AEs after first and second doses of vaccination are summarized in Figure 2 and Table S2. The solicited AEs were mostly mild and the only significant first dose local AE was pain/tenderness in the MVC-COV1901 treatment groups where 69.8% of individuals were affected, compared to 32.8% in the placebo group. The most common systemic AE was malaise/fatigue, which occurred in 30.8% and 24.1% of participants in MVC-COV1901 and placebo groups, respectively.

**Figure 2.**
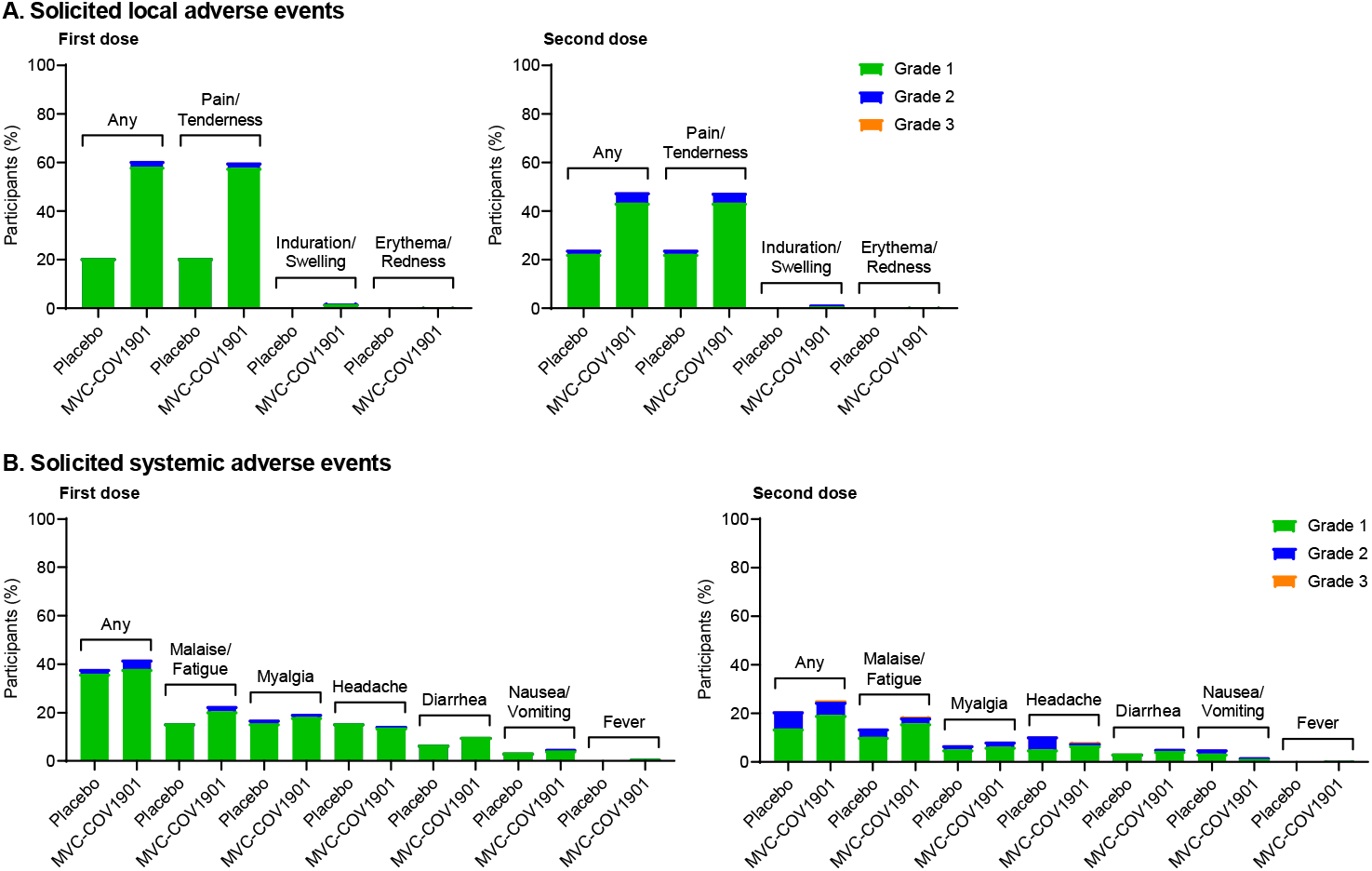
Solicited local (A) and systemic (B) adverse events in adolescents after administration of first or second dose of MVC-COV1901. Events were graded as mild (grade 1), moderate (grade 2), or severe (grade 3)

Except for the absence of fever and a slightly increased incidence of headache, individuals in the placebo group also experienced all systemic AEs but at marginally reduced rates. A total of 26.1% of all participants experienced unsolicited AEs, where the numbers for the placebo group (34.5%) was slightly higher than that for the MVC-COV1901 group (24.6%) (Table S3). Eight individuals presented unsolicited AEs equal or greater than Grade 3 but all were deemed to be unrelated to intervention. No case of SAEs, AESI, VAED, and deaths were reported.

Primary immunogenicity endpoint assessment was performed on Day 57 by comparing GMT and SCR between the adolescent and young adult groups. After conversion to IU/mL, the GMTs of adolescent and young adults were 648.5 [95% CI: 608.6 to 690.9] and 559.5 [95% CI: 512.1 to 611.3], respectively (Table 2). The GMT ratio was 1.16 [95% CI: 1.04 to 1.29], which was higher than the value of 0.67, thus demonstrating non-inferiority of immunogenicity of MVC-COV1901 in adolescents compared to young adults. The SCR based on neutralization assay was 100% in both adolescent and young adult groups (Table 3). Therefore, both groups were similarly completely seroconverted by two doses of MVC-COV1901, again demonstrating the non-inferiority of MVC-COV1901 in the adolescent group.

**Table 2.** Summary of non-inferiority of live SARS-CoV-2 neutralizing antibody titers between adolescent and young adults who received MVC-COV1901 (in IU/mL)

| Per protocol set (PPS)<br>Visit 6 (Day 57) | Adolescents of<br>MVC-COV1901 (N=334) | Young Adults of<br>MVC-COV1901 (N=210) | Ratio adolescent/young<br>adults | P-value |
| --- | --- | --- | --- | --- |
| Median (IQR) | 643.62 (980.4) | 444.87 (498.45) |  |  |
| Q1 ~ Q3 | 429.67 ~ 964.11 | 397.47 ~ 916.71 |  |  |
| Min ~ Max | 136.20 ~ 3466.39 | 120.46 ~ 6976.30 |  |  |
| GMT | 648.47 | 559.54 | 1.16 | P < 0.05* |
| 95% CI | 608.62 ~ 690.93 | 512.05 ~ 611.34 |  |  |

\*Two sample t-test

**Table 3.** Summary of non-inferiority of live SARS-CoV-2 neutralizing antibody seroconversion rate (SCR) between adolescent and young adults who received MVC-COV1901.

| Per protocol set (PPS)<br>Visit 6 (Day 57) | Adolescents of<br>MVC-COV1901 (N=334) | Young Adults of<br>MVC-COV1901 (N=210) | Difference<br>adolescent/young adults | P-value |
| --- | --- | --- | --- | --- |
| SCR, n (%) | 334 (100%) | 210 (100%) | -0.0% | N/A** |
| 95% CI | 98.90 ~ 100.00% | 98.26 ~ 100.00% | -0.00% ~ 0.00% |  |

\* CI calculated using normal approximation for binomial proportion of #
\*\*Chi-square test

For secondary immunogenicity endpoint assessment, only interim results up to Visit 6 are available. The levels of neutralizing antibody and IgG were similarly at baseline level at Visit 2 for MVC-COV1901 and placebo groups (Table S4). After two doses of vaccination, the GMT in MVC-COV1901 group increased to 648.5 [95% CI: 608.6 to 690.9], while the GMT for placebo remained at baseline level, thus yielding a ratio of 115.2 [95% CI: 95.4 to 139.2] between MVC-COV1901 and placebo GMT levels (Table S4). Similar results were seen using anti-spike IgG ELISA assay, in which the MVC-COV1901 group already had a GMT of 136 at Visit 4 that further increased to 1632.0 [95% CI: 1514.9 to 1758.0] at Visit 6, and yielded ratios of 27.8 [95% CI: 24.3 to 31.7] and 254.4 [95% CI: to 387.3] compared to placebo group at Visits 4 and 6, respectively (Table S5). In terms of SCR, MVC-COV1901 group was completely seroconverted by Visit 6 as assessed by neutralization assay or IgG ELISA (Table S6). Notably, after only one dose of MVC-COV1901, 97.3% [95% CI: 95.0 ∼ 98.8] of MVC-COV1901 recipients were already seroconverted by the time they were to receive the second dose (Visit 4) (Table S6).

## Discussion

Here we have investigated two doses of SARS-COV-2 subunit vaccine MVC-COV1901 for safety and immunogenicity in adolescents between 12 and 17 years of age. Study design was largely guided by the results from a prior large phase 2 trial of the vaccine in adults. Thus, we believe that two intramuscular doses of MVC-COV1901 at 28 days apart and immunogenicity analysis 28 days after the second shot on Day 57 were optimal for properly assessing the vaccine in the younger age group studied here.

Safety data of MVC-COV1901 were similar overall to placebo for systemic AEs (Figure 2, Table S2). Only local solicited AEs had greater incidences in the vaccine treatment group compared to placebo, but local solicited AEs were grade 1 except for 5.5% of participants who experienced grade 2 symptoms after any dose of either MVC-COV1901 or placebo (Figure 2, Table S2). The AEs are similar to those observed for adult participants in the phase 2 clinical trial and also similar to the V-Watch vaccine safety monitoring program data collected by the Taiwan Centers of Disease Control, in which pain and fatigue/malaise were the most commonly reported AEs and very few incidences of fever [15, 18]. Overall, therefore, the safety data suggest a highly favorable safety profile for the adolescent age group comparable to the adults.

A consistent picture emerged where immunogenicity was as robust in the adolescent age group as in the larger phase 2 trial. Essentially, the primary study endpoints of live virus neutralizing titers and seroconversion rates were non-inferior when compared with data from the phase 2 trial for young adults (Table S4). Similarly, the secondary endpoints of live virus neutralizing antibody titers and anti-spike IgG titers increased at least two orders of magnitude compared to placebo at 28 days after second dose (Table S5). As antibody titers diminish over time, the availability of full results up to Day 209 will be useful in gauging the degree of waning immunity in adolescents, as compared to our phase 1 extension results with three doses of MVC-COV1901 in adults, which tracked antibody decay up to Day 209 prior to administration of third dose as booster [19]. We have previously performed a *post-hoc* analysis of the phase 2 clinical trial and found a negative association between increasing antibody titers and decreasing age [20]. Thus, unsurprisingly, the adolescent group developed greater neutralizing antibody and IgG titers than that of the young adult group (Tables 2).

Several studies of mRNA COVID-19 vaccines in the adolescents and children were completed and contributed to their approval for use in younger population in the US and Europe. Moderna mRNA-1273 was non-inferior in terms of GMT ratio and seroconversion in the adolescents aged 12 to 17 years versus young adults aged 18 to 25 years, and had an efficacy of 93.3% according to the US CDC definition of COVID-19 with an onset of 14 days after the second dose [21]. In another study with mRNA-1273, two doses of mRNA-1273 invoked neutralization responses against Omicron variant pseudovirus that resulted in numerically superior GMT values in adolescents and children compared to adults [22]. BNT162b2 also demonstrated a higher immune response in the 12 to 15 year old adolescents than in adults and had an observed efficacy of 100% according to the definition of COVID-19 cases with an onset of 7 or more days after the second dose [23]. In all, these external results and our results not only clearly showed that COVID-19 vaccines are suitable for adolescents and children, but that they also generate higher immune responses than in the adults. Thus, the use of vaccines in the young population should help to stop this critical chain of transmission.

Limitations apply to this study. The main limitation is that as this is a phase 2 study where efficacy was not assessed directly, but can only be inferred by established correlates of protection against SARS-CoV-2 [21, 22]. The data presented here make the case for further clinical assessment. Secondly, even though most safety events were grade 1, a small number of grade 3 events were observed that need to be evaluated in a greater study population. Notably, though, no grade 4 events were observed and there were also no cases of AESI and VAED. Other limitations relate to questions regarding the evolution of COVID-19 over the last year that clinical trials are in the process of catching up with. These questions include the duration of the antiviral response over several months after the second dose, the suitability of any given vaccine for current and future virus variants and if there is any additional protection afforded by booster shots. As Taiwan has started administering booster doses for adults including MVC-COV1901, future studies will investigate the suitability of MVC-COV1901 as homologous or heterologous booster after primary vaccination of adolescents.

## Data Availability

All data produced in the present work are contained in the manuscript

## Declaration of Competing Interests

L. T.-C. L., C. E. L., M.-Y. L., H.-Y. C. and C. C. are employees of Medigen Vaccine Biologics Corporation and have received grants from the Taiwan Centers of Disease Control, Ministry of Health and Welfare. L.-M. H. declared no conflict of interest. All authors have reviewed and approved of the final version of the manuscript.

## Acknowledgements

We thank the Institute of Biomedical Sciences, Academia Sinica for performing the neutralization assay. We also thank the members of Medigen Vaccine Biologics Corp. in assisting manuscript editing and revision.

## Author Contribution

Concept and design: C. E. L., H.-Y. C., C. C.

Conducting the clinical trial: L.-M. H., C.-H. C., N.-C. C., B.-F. T., C.-Y., L.

Acquisition, interpretation, and analysis of data: L. T.-C. L., C. E. L., M.-Y. L., H.-Y. C.

Drafting of the manuscript: L. T.-C. L., C. E. L., M.-Y. L.

## Notes

### Clinical Trial

NCT04951388

### Author Declarations

The trial protocol and informed consent form were approved by Taiwan Food and Drug Administration (TFDA) and the ethics committees at the conducting sites: Mackay Memorial Hospital Hsinchu (Hsinchu City), Chang-Gung Memorial Hospital Linkou (New Taipei City), Mackay Memorial Hospital (Taipei City), National Taiwan University Hospital Hsinchu (Hsinchu City), and National Taiwan University Hospital (Taipei City).

