## Supplemental figure and tables for "Safety and immunogenicity of SARS-CoV-2 vaccine MVC-COV1901 in adolescents in Taiwan: A double-blind, randomized, placebo-controlled phase 2 trial"

### Figure S1. Timeline of the study

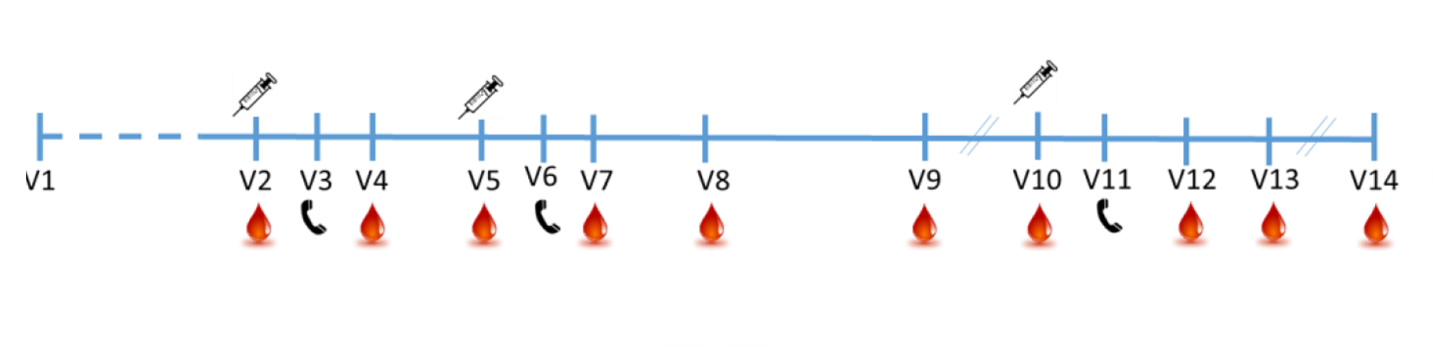

### Table S1. Summary of Demographics and Baseline Characteristics by Adolescent and Young Adults: PPS Subset in CT-COV-22 and Young Adults for Comparison in CT-COV-21

|  | **Adolescents of**  **MVC-COV1901 (N=334)** | **Young Adults of**  **MVC-COV1901 (N=210)** | **Total (N=544)** |
| --- | --- | --- | --- |
| **Age**  Mean (SD)  Min ~ Max | 14.4 (1.65)  12.0 ~ 17.0 | 25.9 (2.74)  20.0 ~ 30.0 | 18.8 (6.02)  12.0 ~ 30.0 |
| **Gender, n (%)** |  |  |  |
| Male | 188 (56.3) | 105 (50.0) | 293 (53.9) |
| Female | 146 (43.7) | 105 (50.0) | 251 (46.1) |
| **Race, n (%)** |  |  |  |
| Asian  Non-Asian | 334 (100.0)  0 | 219 (100.0)  0 | 544 (100.0)  0 |
| **BMI (kg/m^2^),**  Mean (SD)  Min ~ Max | 21.2 (4.2)  15.2 ~ 42.3 | 23.6 (4.1)  14.4 ~ 40.5 | 22.1 (4.3)  14.4 ~ 42.3 |
| **BMI group, n (%)** |  |  |  |
| < 30 kg/m^2^ | 323 (96.7) | 192 (91.4) | 515 (94.7) |
| ≥ 30 kg/m^2^ | 11 (3.3) | 18 (8.6) | 29 (5.3) |
| **HIV antibody, n (%)** | 0 | 1 (0.5) | 1 (0.5) |
| **Pre-vaccination neutralizing antibody against wild type SARS-CoV-2 (≥8), n (%)** | 0 | 0 | 0 |
| **Pre-vaccination anti-SARS-CoV-2 IgG (≥100), n (%)** | 9 (2.7) | 9 (4.3) | 18 (3.3) |
| **Comorbidity, n (%)**  Any  Cardiovascular disease  Cerebrovascular disease  COPD  Liver cirrhosis  Malignancy  HbAlc higher than normal range | 6 (1.8)  0  0  0  0  0  6 (1.8) | 6 (2.9)  1 (0.5)  0  0  0  0  5 (2.4) | 12 (2.2)  1 (0.2)  0  0  0  0  11 (2.0) |

Table S2. Summary of solicited adverse events after each dosing

|  | After any dose | | | After first dose | | | After second dose | | |
| --- | --- | --- | --- | --- | --- | --- | --- | --- | --- |
|  | MVC-COV1901  (N = 341) | Placebo  (N = 58) | Total  (N = 399) | MVC-COV1901  (N = 341) | Placebo  (N = 58) | Total  (N = 399) | MVC-COV1901  (N = 340) | Placebo  (N = 58) | Total  (N = 398) |
| Any AEs | 265 (77.7) | 31 (53.4) | 296 (74.2) | 240 (70.4) | 26 (44.8) | 266 (66.7) | 182 (53.5) | 16 (27.6) | 198 (49.7) |
| Grade 1 | 225 (66.0) | 26 (44.8) | 251 (62.9) | 221 (64.8) | 25 (43.1) | 246 (61.7) | 155 (45.6) | 12 (20.7) | 167 (42.0) |
| Grade 2 | 38 (11.1) | 5 (8.6) | 43 (10.8) | 19 (5.6) | 1 (1.7) | 20 (5.0) | 25 (7.4) | 4 (6.9) | 29 (7.3) |
| Grade 3 | 2 (0.6) | 0 | 2 (0.5) | 0 | 0 | 0 | 2 (0.6) | 0 | 2 (0.5) |
| Any Solicited Local AEs | 238 (69.8) | 19 (32.8) | 257 (64.4) | 207 (60.7) | 12 (20.7) | 219 (54.9) | 163 (47.9) | 14 (24.1) | 177 (44.5) |
| Grade 1 | 217 (63.6) | 18 (31.0) | 235 (58.9) | 199 (58.4) | 12 (20.7) | 211 (52.9) | 148 (43.5) | 13 (22.4) | 161 (40.5) |
| Grade 2 | 21 (6.2) | 1 (1.7) | 22 (5.5) | 8 (2.3) | 0 | 8 (2.0) | 15 (4.4) | 1 (1.7) | 16 (4.0) |
| **Pain/Tenderness** | 238 (69.8) | 19 (32.8) | 257 (64.4) | 205 (60.1) | 12 (20.7) | 217 (54.4) | 162 (47.6) | 14 (24.1) | 176 (44.2) |
| Grade 1 | 218 (63.9) | 18 (31.0) | 236 (59.1) | 197 (57.8) | 12 (20.7) | 209 (52.4) | 148 (43.5) | 13 (22.4) | 161 (40.5) |
| Grade 2 | 20 (5.9) | 1 (1.7) | 21 (5.3) | 8 (2.3) | 0 | 8 (2.0) | 14 (4.1) | 1 (1.7) | 15 (3.8) |
| **Induration/Swelling** | 11 (3.2) | 0 | 11 (2.8) | 7 (2.1) | 0 | 7 (1.8) | 5 (1.5) | 0 | 5 (1.3) |
| Grade 1 | 8 (2.3) | 0 | 8 (2.0) | 6 (1.8) | 0 | 6 (1.5) | 3 (0.9) | 0 | 3 (0.8) |
| Grade 2 | 3 (0.9) | 0 | 3 (0.8) | 1 (0.3) | 0 | 1 (0.3) | 2 (0.6) | 0 | 2 (0.5) |
| **Erythema/Redness** | 4 (1.2) | 0 | 4 (1.0) | 2 (0.6) | 0 | 2 (0.5) | 2 (0.6) | 0 | 2 (0.5) |
| Grade 1 | 4 (1.2) | 0 | 4 (1.0) | 2 (0.6) | 0 | 2 (0.5) | 2 (0.6) | 0 | 2 (0.5) |
| Any Solicited Systemic AEs | 162 (47.5) | 26 (44.8) | 188 (47.1) | 143 (41.9) | 22 (37.9) | 165 (41.4) | 86 (25.3) | 12 (20.7) | 98 (24.6) |
| Grade 1 | 134 (39.3) | 21 (36.2) | 155 (38.8) | 130 (38.1) | 21 (36.2) | 151 (37.8) | 66 (19.4) | 8 (13.8) | 74 (18.6) |
| Grade 2 | 26 (7.6) | 5 (8.6) | 31 (7.8) | 13 (3.8) | 1 (1.7) | 14 (3.5) | 18 (5.3) | 4 (6.9) | 22 (5.5) |
| Grade 3 | 2 (0.6) | 0 | 2 (0.5) | 0 | 0 | 0 | 2 (0.6) | 0 | 2 (0.5) |
| **Malaise/Fatigue** | 105 (30.8) | 14 (24.1) | 119 (29.8) | 78 (22.9) | 9 (15.5) | 87 (21.8) | 63 (18.5) | 8 913.8) | 71 (17.8) |
| Grade 1 | 90 (26.4) | 12 (20.7) | 102 (25.6) | 70 (20.5) | 9 (15.5) | 79 (19.8) | 54 (15.9) | 6 (10.3) | 60 (15.1) |
| Grade 2 | 14 (4.1) | 2 (3.4) | 16 (4.0) | 8 (2.3) | 0 | 8 (2.0) | 8 (2.4) | 2 (3.4) | 10 (2.5) |
| Grade 3 | 1 (0.3) | 0 | 1 (0.3) | 0 | 0 | 0 | 1 (0.3) | 0 | 1 (0.3) |
| **Myalgia** | 81 (23.8) | 13 (22.4) | 94 (23.6) | 66 (19.4) | 10 (17.2) | 76 (19.0) | 27 (7.9) | 6 (10.3) | 33 (8.3) |
| Grade 1 | 71 (20.8) | 11 (19.0) | 82 (20.6) | 62 (18.2) | 9 (15.5) | 71 (17.8) | 23 (6.8) | 3 (5.2) | 26 (6.5) |
| Grade 2 | 10 (2.9) | 2 (3.4) | 12 (3.0) | 4 (1.2) | 1 (1.7) | 5 (1.3) | 3 (0.9) | 3 (5.2) | 6 (1.5) |
| **Headache** | 58 (17.0) | 13 (22.4) | 71 (17.8) | 49 (14.4) | 9 (15.5) | 58 (14.5) | 1 (0.3) | 0 | 1 (0.3) |
| Grade 1 | 53 (15.5) | 10 (17.2) | 63 (15.8) | 47 (13.8) | 9 (15.5) | 56 (14.0) | 28 (8.2) | 4 (6.9) | 32 (8.0) |
| Grade 2 | 4 (1.2) | 3 (5.2) | 7 (1.8) | 2 (0.6) | 0 | 2 (0.5) | 21 (6.2) | 3 (5.2) | 24 (6.0) |
| Grade 3 | 1 (0.3) | 0 | 1 (0.3) | 0 | 0 | 0 | 7 (2.1) | 1 (1.7) | 8 (2.0) |
| **Diarrhoea** | 44 (12.9) | 5 (8.6) | 49 (12.3) | 34 (10.) | 4 (6.9) | 38 (9.5) | 18 (5.3) | 2 (3.4) | 20 (5.0) |
| Grade 1 | 41 (12.0) | 5 (8.6) | 46 (11.5) | 34 (10.) | 4 (6.9) | 38 (9.5) | 15 (4.4) | 2 (3.4) | 17 (4.3) |
| Grade 2 | 3 (0.9) | 0 | 3 (0.8) | 0 | 0 | 0 | 3 (0.9) | 0 | 3 (0.8) |
| **Nausea/Vomiting** | 22 (6.5) | 5 (8.6) | 27 (6.8) | 17 (5.0) | 2 (3.4) | 19 (4.8) | 6 (1.8) | 3 (5.2) | 9 (2.3) |
| Grade 1 | 18 (5.3) | 4 (6.9) | 22 (5.5) | 15 (4.4) | 2 (3.4) | 17 (4.3) | 4 (1.2) | 2 (3.4) | 6 (1.5) |
| Grade 2 | 4 (1.2) | 1 (1.7) | 5 (1.3) | 2 (0.6) | 0 | 2 (0.5) | 2 (0.6) | 1 (1.7) | 3 (0.8) |
| **Fever** | 4 (1.2) | 0 | 4 (1.0) | 3 (0.9) | 0 | 3 (0.8) | 1 (0.3) | 0 | 1 (0.3) |
| Grade 1 | 4 (1.2) | 0 | 4 (1.0) | 3 (0.9) | 0 | 3 (0.8) | 1 (0.3) | 0 | 1 (0.3) |

Table S3. Summary of unsolicited adverse events and other adverse events

|  | After any dose, n (%) | | |
| --- | --- | --- | --- |
|  | MVC-COV1901  (N = 341) | Placebo  (N = 58) | Total  (N = 399) |
| Unsolicited AEs | 84 (24.6) | 20 (34.5) | 104 (26.1) |
| Related unsolicited AEs | 31 (9.1) | 7 (12.1) | 38 (9.5) |
| Unsolicited AEs ≥ Grade 3 | 6 (1.8) | 2 (3.4) | 8 (2.0) |
| Related unsolicited AEs ≥ Grade 3 | 0 | 0 | 0 |
| SAEs | 0 | 0 | 0 |
| Related SAEs | 0 | 0 | 0 |
| AESI | 0 | 0 | 0 |
| VAED | 0 | 0 | 0 |
| AEs leading to study intervention discontinuation | 0 | 0 | 0 |
| AEs leading to study withdrawal | 0 | 0 | 0 |
| Death | 0 | 0 | 0 |

Abbreviations: AE = adverse event; AESI = adverse events of special interest; N = number of participants in the population; n = number of participants with events; SAE = serious adverse event; VAED = vaccine-associated enhanced disease

### Table S4. Summary of GMT and GMT ratio of live SARS-COV-2 neutralizing antibody titer by treatment group (IU/mL)

| **Per protocol set (PPS)**  **Visit 2 (Day 1)** | **MVC-COV1901 (N=334)** | **Placebo (N=46)** | **Ratio MVC/Placebo** | **P-value** |
| --- | --- | --- | --- | --- |
| **Median (IQR)** | 5.04 (0) | 5.04 (0) |  |  |
| **Q1 ~ Q3** | 5.04 ~ 5.04 | 5.04 ~ 5.04 |  |  |
| **Min ~ Max** | 5.04 ~ 5.04 | 5.04 ~ 5.04 |  |  |
| **GMT**  95% CI | 5.04  5.04 ~ 5.04 | 5.04  5.04 ~ 5.04 | 1.00  1.00 ~ 1.00 | N/A |
| **Per protocol set (PPS)**  **Visit 6 (Day 57)** | **MVC-COV1901 (N=334)** | **Placebo (N=46)** | **Ratio MVC/Placebo** | **P-value** |
| **Median (IQR)** | 643.62 (619.59) | 5.04 (0) |  |  |
| **Q1 ~ Q3** | 429.67 ~ 964.11 | 5.04 ~ 5.04 |  |  |
| **Min ~ Max** | 136.20 ~ 3466.39 | 5.04 ~ 787.75 |  |  |
| **GMT**  95% CI | 648.47  608.62 ~ 690.93 | 5.63  4.51 ~ 7.02 | 115.18 | P < 0.001* |

*Two sample t-test

### Table S5. Summary of GMT and GMT ratio of anti-S-2P IgG titer by treatment group (BAU/mL)

| **Per protocol set (PPS)**  **Visit 2 (Day 1)** | **MVC-COV1901 (N=334)** | **Placebo (N=46)** | **Ratio MVC/Placebo** | **P-value** |
| --- | --- | --- | --- | --- |
| **Median (IQR)** | 4.56 (0) | 4.56 (0) |  |  |
| **Q1 ~ Q3** | 4.56 ~ 4.56 | 4.56 ~ 4.56 |  |  |
| **Min ~ Max** | 4.56 ~ 36.02 | 4.56 ~ 22.34 |  |  |
| **GMT**  95% CI | 4.70  4.60 ~ 4.80 | 4.82  4.45 ~ 5.22 | 0.98  0.90 ~ 1.06 | 0.5557* |
| **Per protocol set (PPS)**  **Visit 4 (Day 29)** | **MVC-COV1901 (N=334)** | **Placebo (N=46)** | **Ratio MVC/Placebo** | **P-value** |
| **Median (IQR)** | 148.38 (181.76) | 4.56 (0) |  |  |
| **Q1 ~ Q3** | 71.59 ~ 253.35 | 4.56 ~ 4.56 |  |  |
| **Min ~ Max** | 10.67 ~ 2810.69 | 4.56 ~ 24.44 |  |  |
| **GMT**  95% CI | 136.00  122.85 ~ 150.55 | 4.90  4.49 ~ 5.35 | 27.76  24.30 ~ 31.71 | P < 0.0001* |
| **Per protocol set (PPS)**  **Visit 6 (Day 57)** | **MVC-COV1901 (N=334)** | **Placebo (N=46)** | **Ratio MVC/Placebo** | **P-value** |
| **Median (IQR)** | 1637.72 (1548.30) | 4.56 (0) |  |  |
| **Q1 ~ Q3** | 1024.54 ~ 2572.84 | 4.56 ~ 4.56 |  |  |
| **Min ~ Max** | 281.26 ~ 8530.76 | 4.56 ~ 7640.28 |  |  |
| **GMT**  95% CI | 1631.96  1514.93 ~ 1758.04 | 6.42  4.24 ~ 9.71 | 254.37  167.08 ~ 387.25 | P < 0.0001* |

*Two sample t-test

### Table S6. Seroconversion rate based on neutralizing antibody titers and anti-S-2P IgG titers by treatment group

| **Per protocol set (PPS)**  **Visit 4 (Day 29) (IgG)** | **MVC-COV1901 (N=334)** | **Placebo (N=46)** | **Difference MVC/Placebo** | **P-value** |
| --- | --- | --- | --- | --- |
| **SCR, n (%)**  95% CI | 325 (97.3%)  94.95 ~ 98.76% | 0 (0%) | 97.31%  95.57 ~ 99.04% | P < 0.0001** |
| **Per protocol set (PPS)**  **Visit 6 (Day 57) (Neutralization assay)** | **MVC-COV1901 (N=334)** | **Placebo (N=46)** | **Difference MVC/Placebo** | **P-value** |
| **SCR, n (%)**  95% CI | 334 (100%)  98.90 ~ 100.00% | 1 (2.2%)  0.06 ~ 11.53% | 97.83%  93.61 ~ 102.04% | P < 0.0001** |
| **Per protocol set (PPS)**  **Visit 6 (Day 57) (IgG)** | **MVC-COV1901 (N=334)** | **Placebo (N=46)** | **Difference MVC/Placebo** | **P-value** |
| **SCR, n (%)**  95% CI | 334 (100%)  98.90 ~ 100.00% | 2 (4.3%)  0.53 ~ 14.84% | 95.65%  89.76 ~ 101.55% | P < 0.0001** |

* Exact CI of binomial proportion of #

**Chi-square test
